# Evaluating the relationship between summer season and infant salmonellosis in the United States

**DOI:** 10.1101/2025.05.20.25327655

**Authors:** Kathleen M. Fitzsimmons, Barbara E. Mahon, Matthew P. Fox, Martha M. Werler

## Abstract

Nontyphoidal *Salmonella* infection, or salmonellosis, typically presents with diarrhea, fever, and abdominal cramps and affects over 1 million people in the U.S. annually. Infants and young children are most susceptible to *Salmonella* infection, which can require hospitalization and lead to death. The vast majority of salmonellosis is foodborne and its incidence is highest during hot weather. This study used U.S. age-specific *Salmonella* surveillance data and geographic-specific temperature data to assess the relationship between summer season and *Salmonella* infection according to age group and geographic region. The *Salmonella* infection rate per 100,000 population was highest in infants (111.95), followed by children aged 1-4 (40.66), those aged 5-17 years (12.06), and adults 18 and older (10.84). Compared to winter months (December to February), an excess of approximately 8 infections per 100,000 infants for summer months (June to August) was observed. The corresponding excess was 15 per 100,000 infants for in the South. These findings suggest greater vulnerability among infants to future temperature increases.

## Introduction

Nontyphoidal *Salmonella* infection, or salmonellosis, is an important public health issue in the United States. In 2019, an estimated 1.3 million illnesses, 12,500 hospitalizations, and 238 deaths in the U.S. were due to domestically acquired foodborne Salmonella, and billions of dollars are attributed to *Salmonella* each year (Scallan Walter et al. 2025, CDC 2024). Healthy People 2030 includes reducing *Salmonella* infection as a national food safety objective (USDHHS 2022). Most persons with salmonellosis develop a gastrointestinal illness characterized by diarrhea, fever and abdominal cramps between 12 and 72 hours after infection. While most healthy individuals recover on their own within a few days, invasive infections and severe illness resulting in hospitalization and death may occur, especially among older adults, persons with compromised immune systems and young children (CDC 2024)

Infants (age <1 year) have an incidence rate of *Salmonella* infection approximately eight times that of the general population, and about one out of every four infected infants requires hospitalization (Cheng et al. 2013; Jones et al. 2008). Risk factors for infant infection have not been fully elucidated (Jones et al. 2006; Rowe et al. 2004; Patrick et al. 2010; Jenco 2023). *Salmonella* serotypes have varying transmission pathways and frequencies according to age (Cheng et al. 2013; Crim et al. 2014; Jones et al. 2008; Judd et al. 2019.) While an estimated 94% of all salmonellosis is foodborne, environmental sources (e.g., contact with reptiles; household member with diarrhea in prior four weeks) may be more important for infants (Scallan et al. 2011; Jones et al. 2006; Jenco 2023; Patrick et al. 2010; Schutze et al. 1999; Ackman et al. 2010). Additionally, host factors, such as an immature immune system, may make infants more susceptible to infection and severe illness (Jenco 2023).

Studies, both in the U.S. and abroad, have found that salmonellosis incidence is associated with higher ambient temperature, with peak occurrences during the summer season (Akil et al. 2014; Jiang et al. 2015; Naumova et al. 2007; Uejio 2017; Lake et al. 2009; D’Souza et al 2004; Kovats et al. 2004; Britton et al. 2010; Kendrovski et al. 2011; Lal A et al. 2012). This pattern may be due to temperature directly affecting bacterial growth or temperature-related activities (e.g., food choice, preparation, storage) or other factors (Kovats et al. 2004; Akil et al. 2014). The effect of temperature has also been shown to vary by S*almonella* serotype (Kovats et al. 2004; Lake et al. 2009; Milazzo et al. 2015).

Infants are especially susceptible to salmonellosis, but few studies have examined whether the relationship between ambient temperature and Salmonella infection varies by age and serotype simultaneously. This study sought to assess the relationship between summer season in the U.S. and *Salmonella* infection in infants compared to older age groups, according to geographic region.

## Methods

### Outcome and Exposure

Data on incident human *Salmonella* infections in the U.S. for the years 2010-2015 from the U.S. Centers for Disease Control and Prevention’s Laboratory-based Enteric Disease Surveillance (LEDS) system were used for this study. Laboratory-confirmed human *Salmonella* isolates are reported to LEDS by states. The outcome was defined as laboratory-confirmed nontyphoidal *Salmonella* infection (ODPHP 2025). For exposure, date of specimen collection was used to identify months and seasons. The summer season (exposed) was defined as June, July, and August, which are generally the hottest months in the U.S. December, January, and February constituted the winter season (the unexposed group). For geographic region, states were grouped into four U.S. Census regions (Supplemental Figure 1). For each infection, we evaluated age (< 1 year, i.e., infants; 1-4 years; 5-17 years; and ≥18 years), sex, and geographic region. .

### Population estimates

U.S. Census data files were used to derive population estimates for rate denominators. We obtained estimated numbers of people in each month for April 2010-December 2015 by age in years and sex (“Vintage 2016 Population Estimates: National Monthly Population Estimates” U.S. Census Bureau 2016a) and by age in years, sex, and state (“Vintage 2016 Population Estimates: Characteristics by Single Year of Age”, U.S. Census Bureau 2016b). First, June, July, and August estimates were summed across six years by age group and sex to create the summer group. The same approach for the winter group was used, except for 2010 for which data were not available. We calculated age-specific approximations for 2010 based on 2011-2015 data Population estimates and percentages for U.S. Census regions and divisions were calculated by year, age group, and sex. Then, to estimate monthly and seasonal estimates for each region (or division) by age group, sex, and year, the percentages were multiplied by the corresponding monthly or seasonal totals from the first dataset. They were then combined across years for each region. The assumption with this approach was that, in each respective stratum, the population percent by region did not change by season. This was likely reasonable for all age groups except older adults who might be more likely to travel south in winter. Lastly, for population estimates for the secondary analysis of four infant age groups, the stratum specific totals for all infants were divided by four.

### Analytic Approach

The distribution of infection was examined by age group, and within each age group, by sex and geographic region, and summer versus winter season. Incidence rates were calculated among select subgroups as the number of infections divided by the U.S. resident population, as estimated above, multiplied by 100,000. Incident *Salmonella* infection was compared for summer versus winter months with rate differences (RDs) and 95% confidence intervals. RDs were also calculated within strata of age group, sex and geographic region to assess effect modification.

### Secondary Analyses

#### Age Subgroups of Infants

Prior research has shown differences in salmonellosis by infant age (Cheng et al. 2013). To assess whether the association between salmonellosis incidence and summer versus winter season, we calculated RDs for age subgroups of infants: age < 3 months; 3-5 months; 6-8 months; 9-11 months.

#### Extended Exposure Period

Previous studies have suggested that increases in reported salmonellosis may occur for up to 5 weeks after exposure to high environmental temperature (D’Souza et al. 2004; Kovats et al. 2004; Naumova et al. 2007; Lake et al. 2009). Thus, we also evaluated the potential impact of extending the exposure period one month through September, and the corresponding reference period through March.

## Results

Overall, there were 264,908 nontyphoidal *Salmonella* infections reported to LEDS from 2010 through 2015. Incidence rates decreased with increasing age, with infants having the highest rate with 111.95 infections per 100,000 population, followed by children aged 1-4 (40.66 per 100,000), those aged 5-17 years (12.06), and adults aged 18 and older (10.84). Within sex and geographic region strata, infants consistently had the highest Salmonella infection rates, followed by children aged 1-4 years.

Rate differences for salmonellosis in the summer versus winter by age group and geographic region, as shown in Figure 1, decreased with increasing age and were all greater than zero. Among infants, the rate difference was 7.91 per 100,000 (95% CI: 7.60-8.23), with little evidence of effect modification by sex. The effect of summer versus winter did appear to be modified by geographic region, with smaller risk differences in the West and Midwest and an excess of 15.02 per 100,000 (95% CI: 14.37-15.67) in the South. Among children ages 1-4 years, the risk difference was 3.29 per 100,000 (95% CI: 3.20-3.39), with a similar pattern of effect modification as observed among infants. Rate differences for the older age groups hovered near 1 per 100,000 overall and within strata.

**Figure 1.**
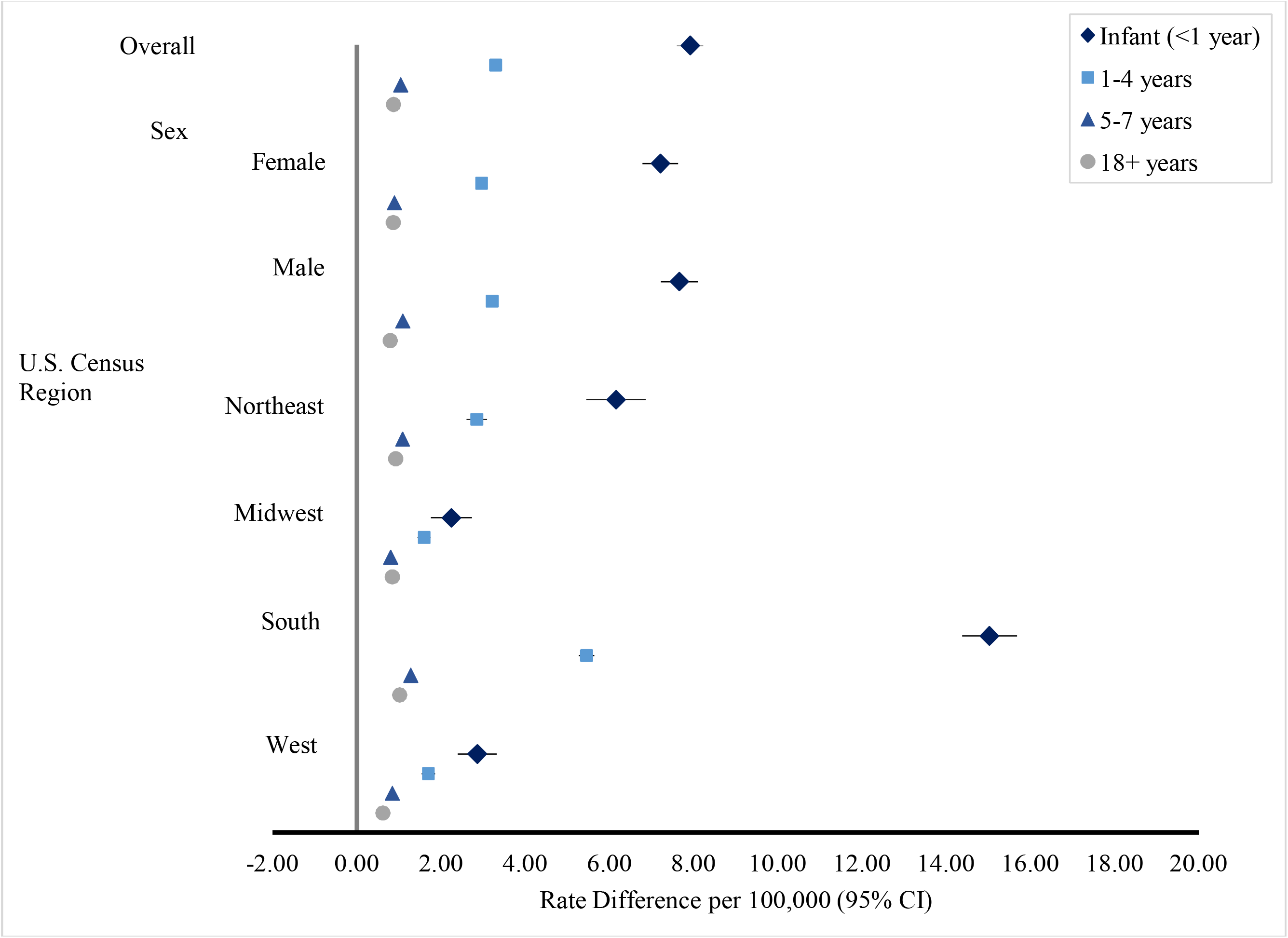
Rate differences and 95% confidence intervals comparing summer and winter incidence of nontyphoidal salmonellosis by age group, sex, and U.S. Census region, 2010-2015.

### Secondary Analyses

Incidence rates of nontyphoidal salmonellosis was highest among infants aged 3-5 months (139.77 per 100,000), followed by those aged 6-8 months (105.34), <3 months (104.89) and 9-11 months (95.29). Across infant age groups, rate differences for summer versus winter were consistently elevated (Figure 2), for which rate differences ranged from 7.26 (95% CI: 6.65-7.87) per 100,000 in 6-8 month olds to 9.32 (95% CI: 8.62-10.03) in 3-5 month olds. Findings from stratified analyses were generally consistent across infant age groups and with those for all infants described above. Overall, the rate difference was highest among infants aged 3-5 months in the South region, who experienced 18.27 (95% CI: 16.79-19.75) excess cases per 100,000 in the Summer compared to Winter.

**Figure 2.**
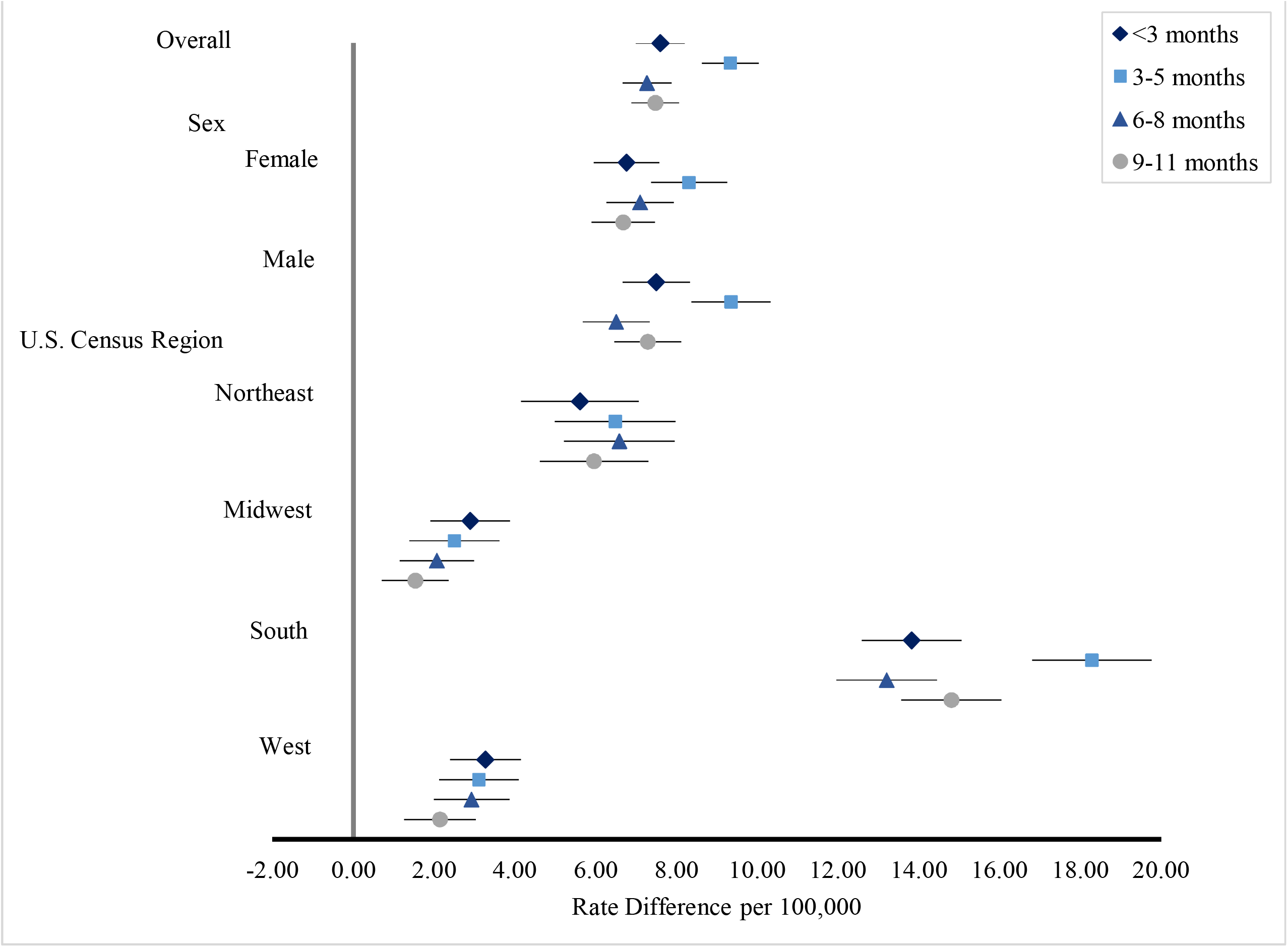
Rate differences and 95% confidence intervals comparing summer and winter incidence of nontyphoidal salmonellosis by infant age group, sex, and U.S. Census region, 2010-2015.

After extending the exposure period through September and the corresponding reference period through March, rate differences increased slightly for infants, did not change for young children aged 1-4 years, and slightly decreased for older children aged 5-17 years and adults (Supplemental Table 4). By geography, infants in the South were most impacted by extending the time frame, where their summer rate difference increased (16.96 per 100,000, 95% CI: 16.38-17.54). Conversely, the effect decreased slightly for infants in the Northeast region and essentially did not change for those in the Midwest or West.

## Discussion

This study used U.S. data on laboratory-confirmed nontyphoidal *Salmonella* infections from 2010-2015 to examine age-specific risk of infection during the summer months and in southern states, when and where average ambient temperatures are highest. As has been reported by others, infants experienced the highest *Salmonella* infection rate; however, this is the first study to examine rates by season and region. The higher rate in infants was especially apparent during June to August and in the South. Compared to December to February, we observed an excess of approximately 8 infections per 100,000 infants for those summer months and 15 per 100,000 infants for summer months in the South. Extending our definition of summer to include September revealed an even higher rate of salmonellosis for infants in the South. These findings suggest greater vulnerability among infants to future temperature increases.

It is well established that incidence of *Salmonella* infection peaks in summer (Akil et al. 2014; Jiang et al. 2015; Naumova et al. 2007; Uejio 2017; Lake et al. 2009; D’Souza et al. 2004; Kovats et al. 2004; Britton et al. 2010; Kendrovski et al. 2011; Lal et al. 2012), however seasonal patterns by age and geographic regions simultaneously are less studied (USDHHS 2022). Our findings are consistent with higher rates in the summer, and are the first to show this pattern in all age groups, in males and females, and in the South, Northeast, West, and Midwest regions of the U.S.

Because incidence and invasiveness of salmonellosis have been shown to vary by infant age, we also explored the effect summer versus winter by age subgroups of infants (Cheng et al. 2013). Findings showed generally similar rate differences among <3 month, 3-5 month, 6-8 month, and 9-11 month age groups, further emphasizing the vulnerability of infants to increasing ambient temperatures.

Prior studies reporting increases in salmonellosis associated with ambient temperature one to five weeks prior to illness onset have attributed the ‘lag’ to possible contamination upstream in the food chain (D’Souza et al. 2004; Kovats et al. 2004; Naumova et al. 2007; Lake et al. 2009). While this might apply to older age groups, it is a less plausible explanation for our findings among infants as contaminated food is likely a less common source of infection in infants and young children, especially the youngest infants. When we extended the exposure period to include September, the excess rate difference in infants in the South were most impacted, whereas we did not observe increases in the effect among infants in the other three census regions. Rather than a delay in cases related to high August temperatures, the observed increased effect among infants in the South might be driven by September temperatures staying high specifically in the South (Supplemental Figure 2). In the three southern states of Mississippi, Tennessee, and Alabama, Akil et al. (2014) found highest numbers of *Salmonella* infections from July through September and a strong positive correlation between infection and monthly temperature.

There are several limitations to note. First, LEDS data likely undercount the true number of salmonellosis cases. Sick persons may not seek medical care, healthcare providers may not order appropriate tests, or the results may not be reported to the health department (CDC 2025). An estimated 29.3 cases of salmonellosis occur for every one that is laboratory confirmed. (Scallan et al. 2011; CDC *Salmonella* Atlas 2014). Importantly, this may vary by age group. For instance, sick infants may be more likely to receive medical care, and in a clinical setting, may be more likely to be tested for *Salmonella* than adults. In this case, the rate for adults may be artificially low. However, as long as the under-recognition and underreporting are consistent throughout the year (i.e., not associated with season/temperature), this should not impact our findings by age group. (Kovats et al. 2004) Furthermore, because LEDS is a passive surveillance system, reporting of confirmed isolates by states varies somewhat year to year. However, Chai et al. (2012) found that from 2004-2009, the annual incidence rates of reported infection with the most common serotype of *Salmonella* (serotype Enteritidis) for the ten FoodNet states were similar for LEDS and FoodNet, which is considered more comprehensive since it conducts active surveillance (Chai et al. 2012).

There were limitations related to our exposure measure. In this study, we used date of specimen collection in lieu of date of exposure or date of illness onset. However, we do not expect this to impact our conclusions since the delay between these dates is likely days and we used three-month time intervals (Kovats et al. 2004). Next, we could not exclude travel-associated cases that were unlikely due to local ambient temperature, but these likely made up only a small proportion of cases (Kendall et al. 2012; Jones et al. 2006). We also could not exclude outbreak-associated cases that might have a different relationship with temperature than sporadic cases (Kovats et al. 2004). However, the vast majority of salmonellosis cases, especially among infants, are sporadic (Olsen et al. 2000; Haddock 1999). Finally, we used a temperature-based definition of season rather than actual temperature measurements as our exposure, which is less precise. However, D’Souza et al. (2004) found that, in the five Australian cities included in their study, average monthly temperature explained the seasonal variation in salmonellosis notifications (D’Souza et al. 2004). Furthermore, temperature has been shown to be a key predictor of salmonellosis. Other meteorological factors like precipitation (Akil et al. 2014) and relative humidity (Kovats et al. 2004; Zhang et al. 2012) are considered less important, although Jiang et al. (2015) reported a positive association between extreme precipitation events and salmonellosis risk in Maryland, especially near the coast.

In conclusion, this study confirmed that infants, especially those in the South, are at increased risk of salmonellosis in summer compared to winter. Findings revealed that, across strata, the absolute effect among infants exceeded that of any other age group and signifies a greater impact on infants and suggests enhanced vulnerability of this group to increases in temperature. Our findings serve as a baseline for future studies examining the potential impact of ambient temperature on *Salmonella* incidence by age and geographic region in the U.S.

**Table 1.** Incidence rates per 100,000 population of nontyphoidal salmonellosis by age group and select characteristics, 2010-2015.

|  | Infant (<1 year) |  |  | 1-4 years |  |  | 5-17 years |  |  | ≥18 years |  |  |
| --- | --- | --- | --- | --- | --- | --- | --- | --- | --- | --- | --- | --- |
|  | No. of Cases | Population | Rate | No. of Cases | Population | Rate | No. of Cases | Population | Rate | No. of Cases | Population | Rate |
| Overall<sup>a</sup> | 26564 | 23728308 | 111.95 | 39166 | 96314118 | 40.66 | 38902 | 322577309 | 12.06 | 156920 | 1.448E+09 | 10.84 |
| Sex |  |  |  |  |  |  |  |  |  |  |  |  |
| Female | 11634 | 11594607 | 100.34 | 17768 | 47108639 | 37.72 | 17076 | 157729158 | 10.83 | 85092 | 743958801 | 11.44 |
| Male | 13420 | 12133702 | 110.6 | 19463 | 49205479 | 39.55 | 20160 | 164848155 | 12.23 | 65257 | 703833624 | 9.27 |
| Census Region |  |  |  |  |  |  |  |  |  |  |  |  |
| Northeast | 3251 | 3815575 | 85.2 | 6087 | 15408054 | 39.51 | 6666 | 53340585 | 12.5 | 29470 | 262575966 | 11.22 |
| Midwest | 2804 | 4996524 | 56.12 | 4627 | 20409833 | 22.67 | 6493 | 69672189 | 9.32 | 33767 | 309494410 | 10.91 |
| South | 17095 | 9072806 | 188.42 | 21724 | 36778822 | 59.07 | 17066 | 121922157 | 14 | 62228 | 539610138 | 11.53 |
| West | 3413 | 5818545 | 58.66 | 6728 | 23640103 | 28.46 | 8677 | 77645020 | 11.18 | 31450 | 336474185 | 9.35 |

<sup>a</sup>Strata totals might not add to Overall total due to missings.

## Data Availability

A data use agreement was completed with the U.S. Centers for Disease Control and Prevention to access the Laboratory-based Enteric Disease Surveillance data.

## Notes

### Competing Interest Statement

The authors have declared no competing interest.

### Funding Statement

This work was completed as part of the requirements of a PhD degree.

### Author Declarations

The Institutional Review Board of the Boston University Medical Center waived ethical approval for this work as non-human subjects research.

