## Supplement for "Evaluating the relationship between summer season and infant salmonellosis in the United States"

**Supplemental Figures 1 and 2**

**Supplemental Tables 1-4**

Supplemental Figure 1.

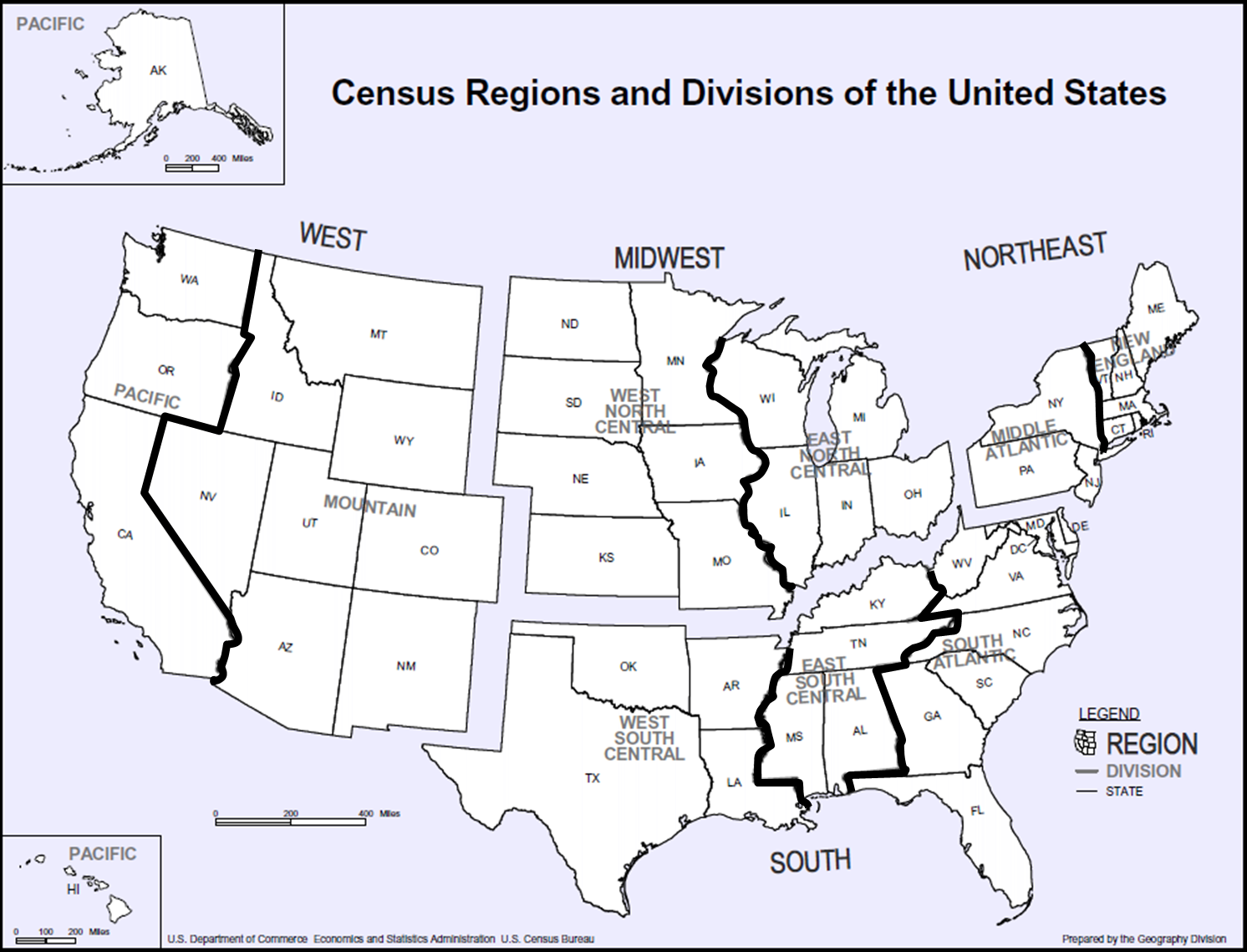

Source: U.S. Census Bureau. Available at https://www2.census.gov/geo/pdfs/maps-data/maps/reference/us_regdiv.pdf, Accessed 57/2025

Supplemental Figure 2.

Average monthly ambient temperature (ºF) across the contiguous United States by month and season, 1981-2010.
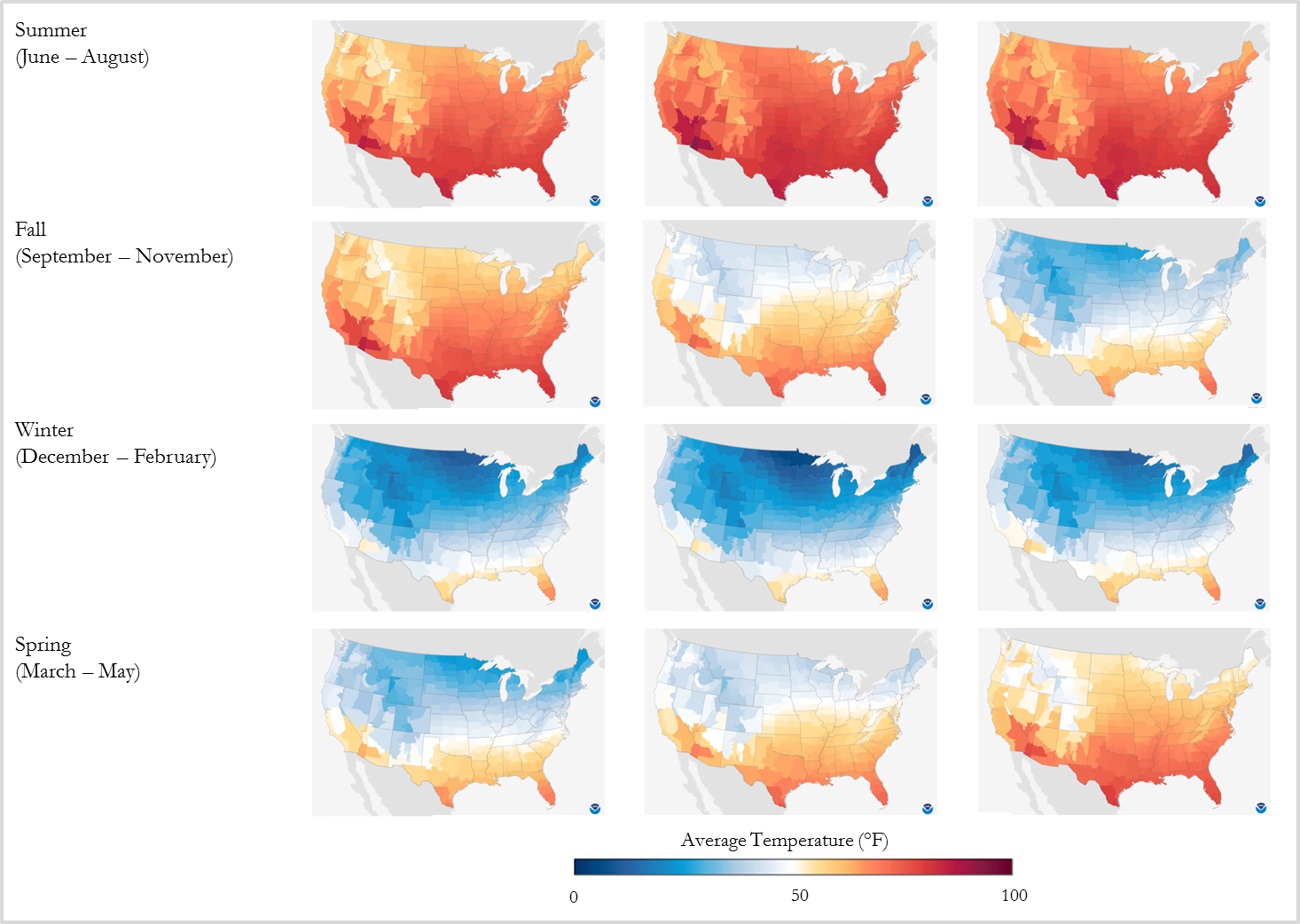

Source: National Oceanic and Atmospheric Administration. Available at. <https://www.ncei.noaa.gov/products/land-based-station/us-climate-normals>. Accessed May 14, 2025

Supplemental Table 1.

Cases of nontyphoidal salmonellosis cases by age group and select characteristics, U.S., Laboratory-based Enteric Disease Surveillance, 2010-2015

|  |  | | Age group | | | | | | | |  | |
| --- | --- | --- | --- | --- | --- | --- | --- | --- | --- | --- | --- | --- |
|  | Total Cases | | Infants (< 1 year) | | 1-4 years | | 5-17 years | | ≥18 years | | Age Missing | |
|  | No. | % | No. | % | No. | % | No. | % | No. | % | No. | % |
| Total | 264908 | 100.0 | 26564 | 100.0 | 39166 | 100.0 | 38902 | 100.0 | 156920 | 100.0 | 3356 | 100.0 |
| Sex |  |  |  |  |  |  |  |  |  |  |  |  |
| Female | 132653 | 50.1 | 11634 | 43.8 | 17768 | 45.4 | 17076 | 43.9 | 85092 | 54.2 | 1083 | 32.3 |
| Male | 119296 | 45.0 | 13420 | 50.5 | 19463 | 49.7 | 20160 | 51.8 | 65257 | 41.6 | 996 | 29.7 |
| Missing | 12959 | 4.9 | 1510 | 5.7 | 1935 | 4.9 | 1666 | 4.3 | 6571 | 4.2 | 1277 | 38.1 |
| Census Region |  |  |  |  |  |  |  |  |  |  |  |  |
| Northeast | 46173 | 17.4 | 3251 | 12.2 | 6087 | 15.5 | 6666 | 17.1 | 29470 | 18.8 | 699 | 20.8 |
| Midwest | 48496 | 18.3 | 2804 | 10.6 | 4627 | 11.8 | 6493 | 16.7 | 33767 | 21.5 | 805 | 24.0 |
| South | 119252 | 45.0 | 17095 | 64.4 | 21724 | 55.5 | 17066 | 43.9 | 62228 | 39.7 | 1139 | 33.9 |
| West | 50981 | 19.2 | 3413 | 12.9 | 6728 | 17.2 | 8677 | 22.3 | 31450 | 20.0 | 713 | 21.3 |
| Missing | 6 | 0.0 | 1 | 0.0 | 0 | 0.0 | 0 | 0.0 | 5 | 0.0 | 0 | 0.0 |

**Supplementary Table 2**. Comparison of incidence rates of nontyphoidal salmonellosis by season, age group, sex, and geography, U.S., 2010-2015.

|  | Summer | | | Winter | | | Comparison | | |
| --- | --- | --- | --- | --- | --- | --- | --- | --- | --- |
|  | No. of cases | Population | Rate | No. of cases | Population | Rate | RD | 95% CI | |
|  | Infants (<1 year) | | | | | | | | |
| Total | 9315 | 71118861 | 13.10 | 3697 | 71275500 | 5.19 | 7.91 | 7.60 | 8.23 |
| Sex |  |  |  |  |  |  |  |  |  |
| Female | 4075 | 34752378 | 11.73 | 1575 | 34828276 | 4.52 | 7.20 | 6.78 | 7.63 |
| Male | 4680 | 36366483 | 12.87 | 1901 | 36447225 | 5.22 | 7.65 | 7.22 | 8.09 |
| Census Region |  |  |  |  |  |  |  |  |  |
| Northeast | 1211 | 11448095 | 10.58 | 508 | 11473309 | 4.43 | 6.15 | 5.44 | 6.86 |
| Midwest | 856 | 14991366 | 5.71 | 521 | 15024384 | 3.47 | 2.24 | 1.76 | 2.73 |
| South | 6152 | 27221676 | 22.60 | 2068 | 27281631 | 7.58 | 15.02 | 14.37 | 15.67 |
| West | 1096 | 17457724 | 6.28 | 599 | 17496175 | 3.42 | 2.85 | 2.39 | 3.32 |
| Age 1-4 years | | | | | | | | | |
| Total | 14659 | 288738075 | 5.08 | 5164 | 289278573 | 1.79 | 3.29 | 3.20 | 3.39 |
| Sex |  |  |  |  |  |  |  |  |  |
| Female | 6601 | 141222780 | 4.67 | 2430 | 141495537 | 1.72 | 2.96 | 2.82 | 3.09 |
| Male | 7270 | 147515295 | 4.93 | 2540 | 147783036 | 1.72 | 3.21 | 3.08 | 3.34 |
| Census Region |  |  |  |  |  |  |  |  |  |
| Northeast | 2260 | 46228587 | 4.89 | 947 | 46315124 | 2.04 | 2.84 | 2.60 | 3.08 |
| Midwest | 1673 | 61235361 | 2.73 | 699 | 61349989 | 1.14 | 1.59 | 1.44 | 1.75 |
| South | 8432 | 110347029 | 7.64 | 2422 | 110553591 | 2.19 | 5.45 | 5.27 | 5.64 |
| West | 2294 | 70927098 | 3.23 | 1096 | 71059869 | 1.54 | 1.69 | 1.53 | 1.85 |

| **Supplemental Table 2 continued** | | | | | | | | | | |
| --- | --- | --- | --- | --- | --- | --- | --- | --- | --- | --- |
|  | Summer | | | | Winter | | | Comparison | | |
|  | | No. of cases | Population | Rate | No. of cases | Population | Rate | RD | 95% CI | |
|  | |  | Age 5-17 years | | | | | | | |
| Total | | 15167 | 967733271 | 1.57 | 5137 | 967710849 | 0.53 | 1.04 | 1.01 | 1.07 |
| Sex | |  |  |  |  |  |  |  |  |  |
| Female | | 6539 | 473201470 | 1.38 | 2344 | 473145102 | 0.50 | 0.89 | 0.85 | 0.93 |
| Male | | 7958 | 494531801 | 1.61 | 2591 | 494565787 | 0.52 | 1.09 | 1.04 | 1.13 |
| Census Region | |  |  |  |  |  |  |  |  |  |
| Northeast | | 2665 | 160020667 | 1.67 | 932 | 160016959 | 0.58 | 1.08 | 1.01 | 1.16 |
| Midwest | | 2563 | 209015145 | 1.23 | 894 | 209010303 | 0.43 | 0.80 | 0.74 | 0.85 |
| South | | 6685 | 365763983 | 1.83 | 2011 | 365755509 | 0.55 | 1.28 | 1.23 | 1.33 |
| West | | 3254 | 232933476 | 1.40 | 1300 | 232928079 | 0.56 | 0.84 | 0.78 | 0.90 |
|  | | Ages ≥18 years | | | | | | | | |
| Total | | 59651 | 4344550368 | 1.37 | 22025 | 4338570235 | 0.51 | 0.87 | 0.85 | 0.88 |
| Sex | |  |  |  |  |  |  |  |  |  |
| Female | | 31602 | 2232460583 | 1.42 | 12397 | 2229528102 | 0.56 | 0.86 | 0.84 | 0.88 |
| Male | | 25365 | 2112089785 | 1.20 | 8747 | 2109042132 | 0.41 | 0.79 | 0.77 | 0.80 |
| Census Region | |  |  |  |  |  |  |  |  |  |
| Northeast | | 11630 | 787743540 | 1.48 | 4398 | 786659202 | 0.56 | 0.92 | 0.89 | 0.95 |
| Midwest | | 12735 | 928501668 | 1.37 | 4948 | 927223574 | 0.53 | 0.84 | 0.81 | 0.87 |
| South | | 24207 | 1618862560 | 1.50 | 7801 | 1616634177 | 0.48 | 1.01 | 0.99 | 1.03 |
| West | | 11078 | 1009442600 | 1.10 | 4878 | 1008053090 | 0.48 | 0.61 | 0.59 | 0.64 |

**Supplemental Table 3.** Incidence rates per 100,000 population of nontyphoidal salmonellosis by infant age group and select characteristics, 2010-2015

|  | Infant age group | | | | | | | | | |
| --- | --- | --- | --- | --- | --- | --- | --- | --- | --- | --- |
|  | 0-3 months | | | | | 3-6 months | | | | |
|  | # cases | Population | Rate | 95% CI | | # cases | Population | Rate | 95% CI | |
|  | 0-3 months | | | | | 3-6 months | | | | |
| Total | 6222 | 5932077 | 104.89 | 102.28 | 107.49 | 8291 | 5932077 | 139.77 | 136.76 | 142.77 |
| Sex |  |  |  |  |  |  |  |  |  |  |
| Female | 2712 | 2898652 | 93.56 | 90.04 | 97.08 | 3571 | 2898652 | 123.20 | 119.15 | 127.24 |
| Male | 3062 | 3033426 | 100.94 | 97.37 | 104.52 | 4276 | 3033426 | 140.96 | 136.74 | 145.19 |
| Census Region |  |  |  |  |  |  |  |  |  |  |
| Northeast | 864 | 953894 | 90.58 | 84.54 | 96.62 | 914 | 953894 | 95.82 | 89.61 | 102.03 |
| Midwest | 710 | 1249131 | 56.84 | 52.66 | 61.02 | 938 | 1249131 | 75.09 | 70.29 | 79.90 |
| South | 3917 | 2268202 | 172.69 | 167.28 | 178.10 | 5421 | 2268202 | 239.00 | 232.64 | 245.36 |
| West | 730 | 1454636 | 50.18 | 46.54 | 53.82 | 1018 | 1454636 | 69.98 | 65.68 | 74.28 |
|  | 6-9 months | | | | | 9-12 months | | | | |
| Total^a^ | 6249 | 5932077 | 105.34 | 102.73 | 107.95 | 5802 | 5932077 | 97.81 | 95.29 | 100.32 |
| Sex |  |  |  |  |  |  |  |  |  |  |
| Female | 2847 | 2898652 | 98.22 | 94.61 | 101.83 | 2504 | 2898652 | 86.38 | 83.00 | 89.77 |
| Male | 3093 | 3033426 | 101.96 | 98.37 | 105.56 | 2989 | 3033426 | 98.54 | 95.00 | 102.07 |
| Census Region |  |  |  |  |  |  |  |  |  |  |
| Northeast | 776 | 953894 | 81.35 | 75.63 | 87.07 | 697 | 953894 | 73.07 | 67.64 | 78.49 |
| Midwest | 643 | 1249131 | 51.48 | 47.50 | 55.45 | 513 | 1249131 | 41.07 | 37.51 | 44.62 |
| South | 3947 | 2268202 | 174.01 | 168.59 | 179.44 | 3810 | 2268202 | 167.97 | 162.64 | 173.31 |
| West | 883 | 1454636 | 60.70 | 56.70 | 64.71 | 782 | 1454636 | 53.76 | 49.99 | 57.53 |

**Supplemental Table 4.** Comparison of incidence rates per 100,000 of nontyphoidal salmonellosis by extended season, age group, sex, and geography, 2010-2015

|  | Summer + September | | | Winter+March | | |  |  |  |
| --- | --- | --- | --- | --- | --- | --- | --- | --- | --- |
|  | Cases | Population | Rate | Cases | Population | Rate | RD | 95% CI | |
|  | Age <1 year | | | | | | | | |
| Total | 12972 | 94840220 | 13.68 | 4866 | 95017820 | 5.12 | 8.56 | 8.28 | 8.83 |
| Sex |  |  |  |  |  |  |  |  |  |
| Female | 5659 | 46343387 | 12.21 | 2066 | 46429726 | 4.45 | 7.76 | 7.39 | 8.13 |
| Male | 6515 | 48496833 | 13.43 | 2529 | 48588096 | 5.20 | 8.23 | 7.84 | 8.61 |
| Census Region |  |  |  |  |  |  |  |  |  |
| Northeast (1) | 1533 | 15268060 | 10.04 | 717 | 15291182 | 4.69 | 5.35 | 4.74 | 5.96 |
| Midwest (2) | 1184 | 19993639 | 5.92 | 708 | 20023917 | 3.54 | 2.39 | 1.96 | 2.81 |
| South (3) | 8800 | 36304921 | 24.24 | 2646 | 36359900 | 7.28 | 16.96 | 16.38 | 17.54 |
| West (4) | 1455 | 23282964 | 6.25 | 794 | 23318223 | 3.41 | 2.84 | 2.45 | 3.24 |
|  | Age 1-4 years | | | | | | | | |
| Total | 19737 | 384955367 | 5.13 | 6910 | 385706605 | 1.79 | 3.34 | 3.25 | 3.42 |
| Sex |  |  |  |  |  |  |  |  |  |
| Female | 8825 | 188281707 | 4.69 | 3218 | 188663743 | 1.71 | 2.98 | 2.87 | 3.10 |
| Male | 9832 | 196673660 | 5.00 | 3425 | 197042862 | 1.74 | 3.26 | 3.15 | 3.38 |
| Census Region |  |  |  |  |  |  |  |  |  |
| Northeast (1) | 2905 | 61632830 | 4.71 | 1321 | 61744701 | 2.14 | 2.57 | 2.37 | 2.78 |
| Midwest (2) | 2179 | 81640146 | 2.67 | 953 | 81788332 | 1.17 | 1.50 | 1.37 | 1.64 |
| South (3) | 11613 | 147116755 | 7.89 | 3143 | 147383789 | 2.13 | 5.76 | 5.60 | 5.92 |
| West (4) | 3040 | 94561355 | 3.21 | 1493 | 94732995 | 1.58 | 1.64 | 1.50 | 1.78 |

| **Supplemental Table 4 continued** | | | |  |  |  |  |  |  |  |
| --- | --- | --- | --- | --- | --- | --- | --- | --- | --- | --- |
|  | Summer + September | | | | Winter+March | | |  |  |  |
|  | Cases | Population | Rate | | Cases | Population | Rate | RD | 95% CI | |
|  | Age 5-17 years | | | | | | | | | |
| Total | 20043 | 1290286074 | 1.55 | | 6959 | 1290309559 | 0.54 | 1.01 | 0.99 | 1.04 |
| Sex |  |  |  | |  |  |  |  |  |  |
| Female | 8635 | 630933910 | 1.37 | | 3181 | 630864586 | 0.50 | 0.86 | 0.83 | 0.90 |
| Male | 10479 | 659352164 | 1.59 | | 3520 | 659445028 | 0.53 | 1.06 | 1.02 | 1.09 |
| Census Region |  |  |  | |  |  |  |  |  |  |
| Northeast (1) | 3479 | 213353934 | 1.63 | | 1311 | 213365075 | 0.61 | 1.02 | 0.95 | 1.08 |
| Midwest (2) | 3236 | 278677776 | 1.16 | | 1227 | 278692329 | 0.44 | 0.72 | 0.67 | 0.77 |
| South (3) | 9121 | 487669414 | 1.87 | | 2638 | 487694880 | 0.54 | 1.33 | 1.29 | 1.37 |
| West (4) | 4207 | 310567844 | 1.35 | | 1783 | 310584062 | 0.57 | 0.78 | 0.73 | 0.83 |
|  | Age ≥18 years | | | | | | | | | |
| Total | 78462 | 5795454582 | 1.35 | | 30119 | 5782080387 | 0.52 | 0.83 | 0.82 | 0.84 |
| Sex |  |  |  | |  |  |  |  |  |  |
| Female | 41496 | 2977941697 | 1.39 | | 16924 | 2971389734 | 0.57 | 0.82 | 0.81 | 0.84 |
| Male | 33460 | 2817512885 | 1.19 | | 11990 | 2810690916 | 0.43 | 0.76 | 0.75 | 0.78 |
| Census Region |  |  |  | |  |  |  |  |  |  |
| Northeast (1) | 14766 | 1051056462 | 1.40 | | 6089 | 1048586468 | 0.58 | 0.82 | 0.80 | 0.85 |
| Midwest (2) | 16696 | 1238864716 | 1.35 | | 6818 | 1235953370 | 0.55 | 0.80 | 0.77 | 0.82 |
| South (3) | 32500 | 2159987189 | 1.50 | | 10504 | 2154911194 | 0.49 | 1.02 | 1.00 | 1.04 |
| West (4) | 14499 | 1346861146 | 1.08 | | 6706 | 1343696007 | 0.50 | 0.58 | 0.56 | 0.60 |
